# ParSE-seq: A Calibrated Multiplexed Assay to Facilitate the Clinical Classification of Putative Splice-altering Variants

**DOI:** 10.1101/2023.09.04.23295019

**Authors:** Matthew J. O’Neill, Tao Yang, Julie Laudeman, Maria Calandranis, Joseph Solus, Dan M. Roden, Andrew M. Glazer

## Abstract

**Background:** Interpreting the clinical significance of putative splice-altering variants outside 2-base pair canonical splice sites remains difficult without functional studies.

**Methods:** We developed Parallel Splice Effect Sequencing (ParSE-seq), a multiplexed minigene-based assay, to test variant effects on RNA splicing quantified by high-throughput sequencing. We studied variants in SCN5A, an arrhythmia-associated gene which encodes the major cardiac voltage-gated sodium channel. We used the computational tool SpliceAI to prioritize exonic and intronic candidate splice variants, and ClinVar to select benign and pathogenic control variants. We generated a pool of 284 barcoded minigene plasmids, transfected them into Human Embryonic Kidney (HEK293) cells and induced pluripotent stem cell-derived cardiomyocytes (iPSC-CMs), sequenced the resulting pools of splicing products, and calibrated the assay to the American College of Medical Genetics and Genomics scheme. Variants were interpreted using the calibrated functional data, and experimental data were compared to SpliceAI predictions. We further studied some splice-altering missense variants by cDNA-based automated patch clamping (APC) in HEK cells and assessed splicing and sodium channel function in CRISPR-edited iPSC-CMs.

**Results:** ParSE-seq revealed the splicing effect of 224 *SCN5A* variants in iPSC-CMs and 244 variants in HEK293 cells. The scores between the cell types were highly correlated (R^2^=0.84). In iPSCs, the assay had concordant scores for 21/22 benign/likely benign and 24/25 pathogenic/likely pathogenic control variants from ClinVar. 43/112 exonic variants and 35/70 intronic variants with determinate scores disrupted splicing. 11 of 42 variants of uncertain significance were reclassified, and 29 of 34 variants with conflicting interpretations were reclassified using the functional data. SpliceAI computational predictions correlated well with experimental data (AUC = 0.96). We identified 20 unique *SCN5A* missense variants that disrupted splicing, and 2 clinically observed splice-altering missense variants of uncertain significance had normal function when tested with the cDNA-based APC assay. A splice-altering intronic variant detected by ParSE-seq, c.1891-5C>G, also disrupted splicing and sodium current when introduced into iPSC-CMs at the endogenous locus by CRISPR editing.

**Conclusions:** ParSE-seq is a calibrated, multiplexed, high-throughput assay to facilitate the classification of candidate splice-altering variants.

## Introduction

Understanding the clinical consequences of genetic variation is one of the greatest challenges in contemporary human genetics^1^. While most research has focused on understanding exonic variants at the protein level, there is an increasing need to explore variant effects on splicing^2-4^. Sequencing studies have estimated that approximately 10% of pathogenic variants may act through disruptions of splicing outside of the canonical splice acceptor (AG) or donor (GT) splice sites, known as cryptic splicing (Figure 1A)^5,6^. Despite these estimates, both intronic and exonic splice-altering variants have been less well-studied compared to protein-coding missense variants, largely due to the difficulty of identifying and functionally investigating this class of variants. Several assays exist for assessing variant-associated splicing, including direct measurements from patient tissue^7-9^, minigene assays^10-12^, and genome-editing at the endogenous locus^13-15^. However, these assays are often not calibrated with large numbers of control benign and pathogenic variants and are too low-throughput to decipher the thousands of potential splice-altering variants in Mendelian disease genes. An alternative approach is high-throughput multiplexed assays that use deep sequencing to read out pools of splicing outcomes. Several successful implementations of this approach have enabled splicing investigation through saturation mutagenesis of specific genes^16,17^ and medium to large variant libraries by pooled oligonucleotide synthesis^6,18-21^. Building upon these seminal contributions, we sought to develop a versatile, accurate, multiplexed assay of putative splice-altering variants that could facilitate clinical classification of variants^2^.

**Figure 1.**
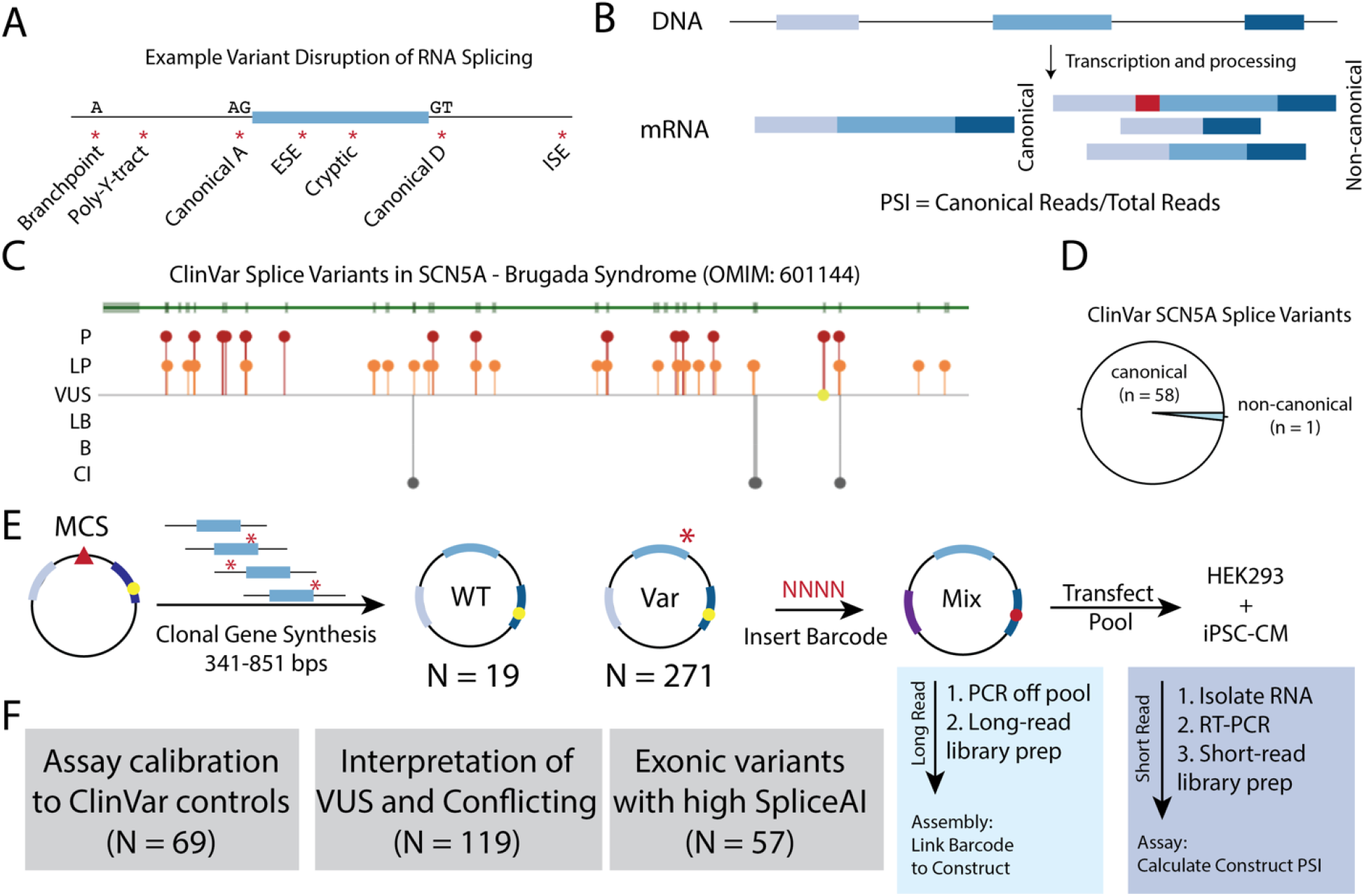
Splice-altering Variants and Assay Schematic. A) Splicing regulatory sequences disrupted or introduced by *cis*-genetic variants. A – acceptor site. D – donor site. ESE – exonic splicing enhancer. ISE – intronic splicing enhancer. Y – pyrimidine. B) Quantification of Percent Spliced In (PSI) from transcripts associated with WT- or variant-containing transcripts. Canonical reads are divided by the total amount of reads for a given exon triplet cassette. C) ClinVar view of splice-altering variants for the gene *SCN5A*. Most known splice variants are P/LP. D) Most *SCN5A* splice variants in ClinVar are associated with canonical splice sites, with only 1 ClinVar example of a non-canonical splice variant. E) Schematic of ParSE-seq assay. A clonal gene library is cloned into a minigene vector (modified pET01), pooled, and barcoded. Barcode are assigned to the WT or variant inserts by long-read sequencing through assembly, and the splicing outcomes are determined with short-read sequencing after transfection into cells. Yellow circle represents restriction site for barcode in downstream exon. F)The *SCN5A* variant library was designed to evaluate three categories of variants: assay calibration, classification of clinically relevant variants, and testing of exonic variants with high SpliceAI scores. VUS – Variant of Uncertain Significance. CI – Conflicting Interpretation.

In the 2015 American College of Medical Genetics and Genomics (ACMG) classification guidelines, “well-validated” functional data could be implemented at the strong level for abnormal assay results (PS3) or normal assay results (BS3).^22^ In 2020, the ClinGen Sequence Variant Interpretation Working Group developed a quantitative approach to “calibrate” such functional assays. The working group recommended that functional assays be deployed on many control benign/likely benign (B/LB) and pathogenic/likely pathogenic (P/LP) variants. These results are used to calculate an odds of pathogenicity (“OddsPath”), the likelihood ratio of variant pathogenicity given normal or abnormal assay results^23,24^. The OddsPath value can be converted into a quantitative evidence strength in the ACMG classification scheme, ranging from supporting to very strong evidence, depending on the number of tested control variants and the concordance of the assay results. Several high-throughput assays of protein-coding variants have been calibrated using this scheme,^25-27^ but to our knowledge, functional assays of splice-altering variants have thus far not. A clinically calibrated, high-throughput splicing assay would facilitate the reclassification of Variants of Uncertain Significance (VUS) and Conflicting Interpretation (CI) variants in disease-associated genes.

*SCN5A* encodes the main voltage-gated sodium channel in the heart, Na_V_1.5. Although Brugada Syndrome has a strong polygenic influence^28^, loss-of-function variants in *SCN5A* are the leading monogenic cause of the arrhythmia disorder Brugada Syndrome (BrS, MIM #601144),^29^ and are also associated with other cardiac disorders^30^. The majority of BrS-associated *SCN5A* variants reported to date are missense, frameshift, indels, or “canonical” splice site variants (variants in 2-bp exon-adjacent splice sites)^31^. We and others have previously deployed low-throughput functional assays to demonstrate splicing defects in several *SCN5A* exonic and intronic variants outside the 2-bp splice sequences^14,32,33^. Enabling larger scale investigations into splice-altering variants in *SCN5A* would decrease clinical uncertainty when managing patients at risk for this potentially fatal arrhythmia syndrome.

In this work, we present a multiplexed splicing assay, Parallel Splice Effect-sequencing (ParSE-seq), to determine the splice-altering consequences of hundreds of intronic and exonic variants in *SCN5A*. We implement ParSE-seq for 244 *SCN5A* variants in Human Embryonic Kidney (HEK293) cells and 224 variants in induced pluripotent-derived cardiomyocyte cells (iPSC-CM). We calibrate the assay with nearly 50 ClinVar-annotated benign and pathogenic variants and compare our experimental outcomes to the *in silico* tool SpliceAI. Using our calibrated strength of evidence, we propose reclassifications for 9 VUS, and contribute new functional data to help adjudicate 29 conflicting interpretation variants in ClinVar. Furthermore, we demonstrate that some missense variants may be incorrectly described as having normal function by conventional cDNA-based patch-clamping assays that cannot assess splicing outcomes.

## Methods

### Selection of Variants

Variants were selected from 3 categories (Figure 1E) – 1) Benign/likely benign (B/LB) and pathogenic/likely pathogenic (P/LP) ClinVar variants to calibrate the assay; 2) VUS and CI variants in *SCN5A* that could be reclassified with splicing functional data; 3) exonic missense and synonymous variants with a strong prediction of splice-disruption (aggregate SpliceAI^5^ score greater than 0.8). Control B/LB and P/LP variants were chosen from ClinVar, accessed on October 2, 2022. We excluded B/LB variants with gnomAD allele count of 4 or less (approximate minor allele frequency <2.5x10^−5^), due to the possibility that such very rare variants may nevertheless be associated with Brugada syndrome^34,35^. We used gnomAD v3.1.2 accessed on October 6^th^ 2022, with Ensembl ID ENST00000333535.9. All selected variants were within 170 base pairs of an exon/intron junction. We selected pathogenic or likely pathogenic variants that were annotated in ClinVar to act through aberrant splicing rather than by altering protein function. The assay requires an acceptor and donor splice site on each end of the insert plasmid’s exon, and is therefore incompatible with the first or last coding exons (2 and 28). In addition, *SCN5A* uses 2 instances of non-canonical AC/AT splice sites between exons 3 and 4, and exons 25 and 26. Therefore, we did not study variants in these exons or in adjacent intronic locations. Furthermore, we were unable to include plasmids with exon 15 due to synthesis incompatibility (high GC content) and exon 17 due to overlap of restriction enzymes used for barcoding.

### Construction of Plasmid Library

The exon trapping vector pET01 (MoBiTec) was mutagenized to introduce two restriction sites (AscI and MfeI) in the 3’ rat insulin exon to produce pAG424 (Supplemental Figure I)^36^. We used an inverse PCR mutagenesis method with primers ag491 and ag492 to introduce the restriction sites which were then verified by Sanger sequencing (all primers are presented in Supplemental Table I)^36^. 290 “clonal genes” containing an exon surrounded by 100-250 bp of intronic sequence on each side were directly synthesized, cloned into pAG424, and sequence verified by Twist Biosciences (South San Francisco, CA). Plasmids containing exons 6, 10, and 12 were shortened from 250 bp due to closely adjacent exons (exon 6) or high GC content at distant intronic sequence which was incompatible with Twist synthesis (exons 10 and 12). We studied the adult form of *SCN5A* exon 6 (exon 6B). All construct sizes are presented in Supplemental Table II. We resuspended each plasmid in water to a concentration of 25 ng/μl. We pooled 50 ng of each plasmid to form a library pool. We also added two additional plasmids (V291 and V304 – Supplemental Table III) that failed Twist synthesis using PCR amplification of the wildtype insert from genomic DNA and QuikChange mutagenesis as previously described^14^.

### Adding a Barcode to the Plasmid Library

We digested the plasmid pool with AscI (NEB) and MfeI (NEB) followed by incubation with Calf Intestinal Phosphatase (NEB). A cloning insert containing an 18-mer barcode was produced. Briefly, ag1371 and ag1372 were annealed, followed by extension to make fully double stranded DNA using Klenow polymerase (NEB)^37^. Due to its small size, the double stranded DNA was then phenol/chloroform extracted and digested using AscI and MfeI (NEB), and was again purified by phenol/chloroform extraction. The pool of minigene plasmids was also digested with AscI and MfeI and cleaned by gel extraction (QIAGEN). The digested vector pool and barcode insert were ligated using T4 ligase (NEB). The ligation product was PCR purified (QIAGEN) and electroporated into ElectroMax DH10B cells (ThermoFisher) using a Gene Pulser Electroporator (BioRad; 2.0 kV, 25 μF, 200 Ω). The resulting bacterial culture was then grown overnight, and DNA was isolated by a maxiprep (QIAGEN) to yield the barcoded plasmid library. Barcode diversity was estimated by plating dilutions of the library on ampicillin plates and counting colonies.

### Assembly Steps

We established the relationship between barcodes and inserts (wildtype [WT] or variant exon and introns) using PacBio long-read SMRT sequencing^38^. We used the pooled barcoded library as template for a PCR with primers mo37 and ag489 using Q5 polymerase (NEB) per the manufacturer’s protocol. Amplification was split among 8 individual reactions and cycle number was minimized to reduce PCR-mediated recombination. The PCR protocol was with 1 denaturation step of 98°C for 30 s, 20 cycles of 98°C 10 s, 55°C for 15 s, and 72°C 1 min, followed by a final 72°C hold for 5 minutes^39^. The reactions were pooled, and used to generate a SMRT Bell 3.0 library (PacBio) according to the manufacturer’s instructions. The library was sequenced for 30 hours with PacBio Sequel II 8M SMRT Cell by Maryland Genomics.

### Computational Pipeline – Assembly

The full computational pipeline is diagrammed in Supplemental Figure II. FASTQ files from the PacBio sequencing were filtered for reads containing barcodes with the predicted 8-bp “prefix” and 6-bp “suffix” sequences immediately adjacent to the 18-bp barcode using Unix and Python. Unique barcodes were identified, and then reads were parsed by barcode into new FASTQ files. For each barcode, the Unix *grep* command was used to count the number of reads containing each designed insert. To be included in the assembly, a barcode was required to be present in at least 50 reads in the assembly sequencing. The barcode identity was assigned as the most frequently aligned insert if that insert represented more than 50% of the read counts. After implementation of these quality control cutoffs, 284 of the 290 targeted plasmids were successfully detected in the plasmid pool.

### Splicing Assay and Preparation of Illumina Library

HEK cells were grown in “HEK media”: Dulbecco’s Eagle’s medium supplemented with 10% fetal bovine serum, 1% non-essential amino acids, and 1% penicillin/streptomycin. For the HEK cell assay, 3 wells of a 6 well plate with HEK cells at 30-40% confluency were independently transfected with the barcoded plasmid library using FuGENE 6 (Promega) per manufacturer’s instructions. For iPSC-CM transfections, we grew and maintained the ‘C2’ line as previously described^40^. iPSCs were differentiated into a monolayer of iPSC-CMs using a chemical differentiation method^41^. Following 30 days of differentiation, iPSC-CMs were dissociated with TrypLE™ Select (ThermoFisher) and replated as single cells. Cells were cultured for an additional 2 days, after which the pooled library was transfected using Viafect (Promega)^42^. For both HEK293 and iPSC-CM experiments, RNA was isolated 24 hours post-transfection using the RNeasy Plus Mini kit (QIAGEN) and reverse transcribed using the primer mo38 (which targets a constant region of the transcribed minigene) with SuperScript III (Invitrogen). Libraries were prepared for Illumina sequencing by touchdown PCR using Q5 polymerase (NEB) with primers binding reference minigene exons, sequence containing Illumina i7 and i5 indexing sequences and Illumina NovaSeq dual indexing motifs (Supplemental Table I). The PCR protocol included a single denaturation step of 98°C for 30 s, touchdown with 10 cycles of 98°C for 10s, 10 cycles of 65°C to 55 °C (decreasing by 1°C per cycle for 15 s, followed by extension at 72 °C for 1 min), followed by an additional 20 cycles of 98°C 10 s, 55°C for 15 s, and 72°C 1 min), followed by a final extension of 72°C for 5 min. PCR amplicons were purified with a PCR purification kit (QIAGEN). Libraries were then sequenced using Illumina NovaSeq paired-end 150 base sequencing to ∼50M reads/sample.

### Computational Pipeline – Assay

A diagram of the computational pipeline is presented in Supplemental Figure II. Reads were filtered for correct barcode prefix and suffix sequences and were divided into separate files by barcode as described above. In all splicing assay studies, each barcode was required to be present in at least 25 reads in each replicate to be included. The percent spliced in (PSI) metric was conducted using *grep* searches for splice junctions corresponding to the WT exon splicing to the reference exons in the R1 and R2 reads:

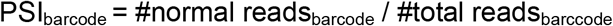

computationally implemented as:

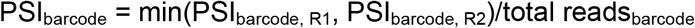

For variants that would alter the coding sequence of the WT exon, a bespoke R1 or R2 junction was created and used for those specific variants. PSIs were then averaged across barcodes for each variant, using the barcode-variant lookup table from the assembly step described above. We used the assigned PSI as the average PSI for each variant across 3 independent transfections into HEK or iPSC-CMs. For each variant, a ΔPSI value was calculated:

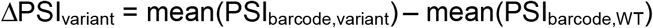

A normalized ΔPSI was then calculated:

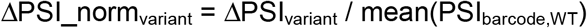

### Statistical Analysis and Data Availability

Statistical analyses were performed in R. Comparison of WT and variant PSI (using mean of three replicate samples) used a 2-sided t-test implemented in R. For the set of studied variants, false discovery rates (FDR) were calculated using the R command *p*.*adjust*. A ΔPSI_norm value of -100% indicates a complete loss of normal splicing, whereas a value of 0 indicates identical splicing to wildtype. Variants with FDR < 0.1 and ΔPSI_norm < -50% were considered splice-altering. Variants with FDR > 0.1 and ΔPSI_norm >= -20% were considered non-splice altering. All other variants were labeled indeterminate in the assay. We excluded variants from our analysis if the standard error of the PSI among the 3 replicates was >0.15. Violin plots, loess best fit curves, and barplots were plotted in R using ggplot2 (see GitHub for code).

### ACMG OddsPath Calculation

We performed assay calibration according to the framework developed by Brnich et al^24^ and extended by Fayer et al^25^. For the high-throughput splicing assay, we calculate the likelihood ratio of pathogenicity (termed OddsPath) for both splice-altering (pathogenic) and non-splice-altering (benign) assay results. First, we removed all variants with indeterminate scores. Then, we calculated Benign and Pathogenic OddsPath values using the equations:

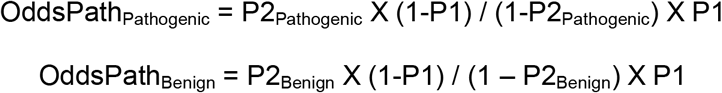

Where, the Prior P1 was defined as:

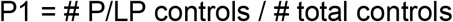

The benign and pathogenic posterior P2 was then calculated:

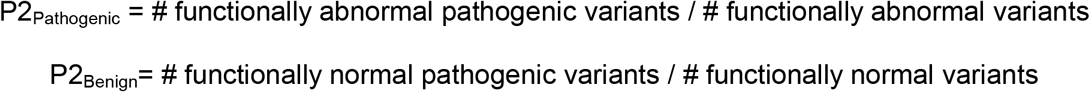

Following the approach recommended by Brnich et al^24^, if P2_Pathogenic_ = 1 it was conservatively estimated to have 1 additional discordant variant (a functionally abnormal benign variant) by the following equation:

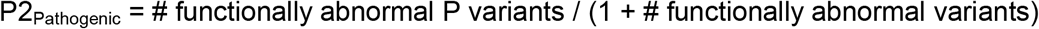

Similarly, if P2_Benign_ = 0, an additional discordant variant was included (a functionally normal pathogenic variant):

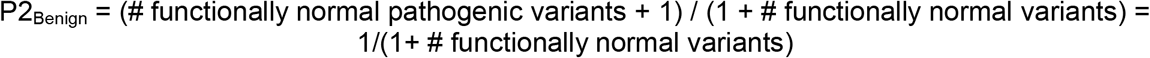

Each posterior was combined with the prior to derive an OddsPath and assign evidence for PS3 and BS3 criteria, respectively.

An interactive Excel worksheet that implements this computation is presented in Supplemental File I.

### In silico predictors

SpliceAI was implemented on the Unix command line using an input VCF file consisting of Human Genome Variation Society GRCh38 coordinates for all variant positions in the splicing library^5^. Pre-computed SpliceAI scores for all possible *SCN5A* variants were downloaded from the SpliceAI dataset hosted at Illumina BaseSpace. We computed an aggregate SpliceAI score to incorporate contributions from each of the 4 predicted categories using a previously described formula^45^:

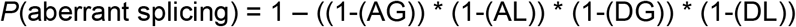

Where AG=probability of acceptor gain, AL=probability of acceptor loss, DG=probability of donor gain, and DL=probability of donor loss. These aggregate scores were used in the analysis of SpliceAI correlations with experimental ΔPSI scores, as well as to select exonic variants above a threshold of 0.8 (high prediction of altered splicing). Receiver Operator Characteristic curves were plotted in R using the library pROC, comparing aggregate SpliceAI scores for the binary outcome of a normal or abnormal splice assay result, removing indeterminate assay results^43^.

REVEL scores^44^ were obtained from https://sites.google.com/site/revelgenomics/. *SCN5A* Bayes structural penetrance^45,46^ scores were obtained from https://variantbrowser.org/.

### ACMG Variant Classification

We determined clinical classifications of variants using the ACMG criteria using an online tool hosted by the University of Maryland (https://www.medschool.umaryland.edu/Genetic_Variant_Interpretation_Tool1.html/)^47^. We implemented functional evidence at the calibrated strength of evidence based on the results of the calibration described above (PS3 strong and BS3 strong). We applied PP3 for an aggregate SpliceAI >0.5, and BP3 for SpliceAI <0.2. We applied PM2 for variants with an allele frequency of < 2.5x10^−5^ per recommendations for BrS^34^.

### High-throughput Electrophysiology

We used an established workflow for generating automated patch clamp data for variants in HEK293T landing pad cells (gift of Kenneth Matreyek).^48-50^ Briefly, we cloned SCN5A variants into cDNA-containing plasmids using Quikchange mutagenesis (Agilent), integrated these plasmids into “landing pad” HEK293 cells^48,49^, used negative and positive selection with iCasp and blasticidin to generate stable lines, and studied the stable lines on the SyncroPatch 384PE automated patch clamping platform^51^ (Nanion). The cloning, patch clamping, and data analysis process has been extensively described by our group in previous publications^50-52^.

### iPSC-CM Maintenance and Differentiation

iPSCs were cultured at 37°C in a humidified 95% air/5% CO_2_ incubator on Matrigel coated plates (BD BioSciences). Cells were maintained in mTeSR plus media (STEMCELL) and passaged every 3-4 days. Stem cell experiments were carried out in the ‘C2’ line previously characterized by our laboratory^40^. At 60-80% confluency, stem cells began differentiation to cardiomyocytes using a previously described chemical method^41,53^. RNA analyses and patch clamp studies were performed at days 35-40 of differentiation. Nonsense-mediated decay inhibition assays were carried out by exposing cells to 100 μM cycloheximide (Sigma-Aldrich) or 1% dimethyl sulfoxide (Sigma-Aldrich) as a vehicle control for 6 hours, followed by harvesting of RNA as described below^54^.

### CRISPR-Cas9 Gene editing

We designed CRISPR-Cas9 guide plasmids based on the 20-bp NGG SpCas9 editor using the online tool CRISPOR^55^. PAM sites were altered in the repair template to a synonymous amino acid for exonic variants and an intronic SNV when intronic; in either case, the disrupting variant was chosen to have a predicted minimal impact on splicing (aggregate SpliceAI <0.05). CRISPR guides were cloned into (pX458)^20^ plasmid (Addgene #48138, a gift of Feng Zhang) as previously described^14^. The cloned guide plasmid and a 151-nucleotide repair template bearing the desired change and PAM site variant were co-electroporated into dissociated iPSCs using the Neon Transfection System (ThermoFisher MPK5000). After 72 hours, cells were dissociated, singularized, and sorted for GFP+ cells using a BD Fortessa 5-laser instrument. Single colonies were picked, DNA extracted using QuickExtract (Lucigen), PCR amplified using primers mo198 and mo199, and Sanger sequenced to identify a colony with a heterozygous edit.

### RNA-seq and Visualization of Splice Junctions

RNA was isolated from iPSC-CMs between day 30 and 35 of differentiation using the RNeasy Minikit (QAIGEN). Integrity of RNA was determined on a 2100 Bioanalyzer (Agilent). The Vanderbilt Technologies for Advanced Genomics core prepared a polyA-selection library and sequenced the library to a depth of 50 million PE100 reads on an Illumina Nova-seq sequencer. FASTQ files were analyzed with fastp^56^, reads were aligned with hisat2^57^, bam files indexed with SAMtools^58^, and quantified in Integrative Genomics Viewer^59^.

### Whole-cell voltage clamp recording of sodium current

Whole-cell voltage clamp experiments in iPSC-CMs were conducted at room temperature (22-23°C). Glass microelectrodes were heat polished to tip resistances of 0.5-1 M_Ω_. Data acquisition was carried out using MultiClamp 700B patch-clamp amplifier and pCLAMP 10 software suite (Molecular Device Corp., San Jose, CA, USA). To record sodium currents in iPSC-CMs, currents were filtered at 5 kHz (−3 dB, four-pole Bessel filter) and digitized at frequency of 2 to 20 kHz by using an analog-to-digital interface (DigiData 1550B, Molecular Device Corp., San Jose, CA, USA). To minimize capacitive transients, capacitance and series resistance were corrected ∼80%. Voltage-clamp protocols used are shown on the figures. Electrophysiological data were analyzed using Clampfit 10 software and the figures were prepared by using Graphing & Analysis software OriginPro 8.5.1 (OriginLab Corp., Northampton, MA, USA). To provide better voltage control of sodium current, the external solution was K^+^-free and Ca^2+^-free with a lower sodium concentration (50 mmol/L), containing (in mM) NaCl 50, NMDG 85, glucose 10, HEPES 10, adjusted to pH 7.4 by NaOH. The pipette (intracellular) solution had (in mM) NaF 5, CsF 110, CsCl 20, EGTA 10, and HEPES 10, adjusted to pH 7.3 by CsOH. To eliminate the overlapped L-/T-type inward calcium currents and outward potassium currents, different blockers (1 μM nisoldipine, 200 μM NiCl_2_, and 200 μM 4-aminopyride) were added into the cell bath solution. To record sodium current, cells were held at -100 mV and current was elicited with a 50-ms pulse from -100 to +40 mV in 10 mV increments. Current densities were expressed in the unit of pA/pF after normalization to cell size (pF), generated from the cell capacitance calculated by the function of Membrane Test (OUT 0) in pCLAMP 10 software.

## Results

### Methodology Overview and Feasibility

We envisioned a high-throughput minigene-based assay that would build upon labor-intensive single-variant minigene studies and extend previous high-throughput functional assays by establishing a platform to investigate variants for higher clinical relevance. Variants disrupt RNA splicing through disruption or creation of splice sites or splicing regulatory elements^3^ (Figure 1A). Our assay used a minigene plasmid containing a test construct bearing exon and surrounding native intronic sequence, flanked by rat insulin exons 1 and 2. When feasible, we included large (250 bp) native intronic flanking regions on either side of the exon. We barcoded and pooled these minigene plasmids and used high-throughput RNA-sequencing to quantify Percent Spliced In (PSI) of the wildtype (WT) or variant test exons (Figure 1B). We designed a library to test the splicing effects of variants in *SCN5A*. Most ClinVar-annotated splice-altering variants in *SCN5A* are at the canonical splice sites, motivating us to explore other variants that may affect splicing in this gene (Figures 1C and D). Towards this goal, we introduced two restriction sites into the 3’ exon of the minigene vector pET01 to allow for a barcode to be included in the spliced RNA-sequencing read^14^ (Figure 1E; see Methods for full details). This design enabled the linkage of reads to variants even if the variant was not included in the spliced transcript.

### Assay Implementation

We designed a library of 290 WT and variant plasmids to test three questions (Figure 1F) – 1) How well does the assay perform on ClinVar P/LP and B/LB control variants? 2) Can these results help reclassify VUS or resolve variants with conflicting interpretations (CI)? 3) What is the prospective accuracy of SpliceAI for exonic synonymous and missense variants? Accordingly, we adopted a previously published minigene to incorporate a site for barcoding to enable multiplexed experiments (Figure 2A). We would first link barcodes to wildtype or variant-containing plasmids, and then use barcode sequencing to quantify percent spliced in for each plasmid (PSI; Figure 2B). After introducing an 18bp poly-N barcode, we used long read sequencing of the plasmid pool to link each barcode to the insert (assembly). We identified 3,238 unique barcodes linked to 284 unique inserts (290 possible; median 11 barcodes per insert; Figure 2C). Comprehensive library characteristics are presented in Supplemental Table III. Barcode frequencies were highly correlated when quantified by PacBio or Illumina sequencing (*R* = 0.82, p-value = 4.0*10^-125;^ Supplemental Figure III).

**Figure 2.**
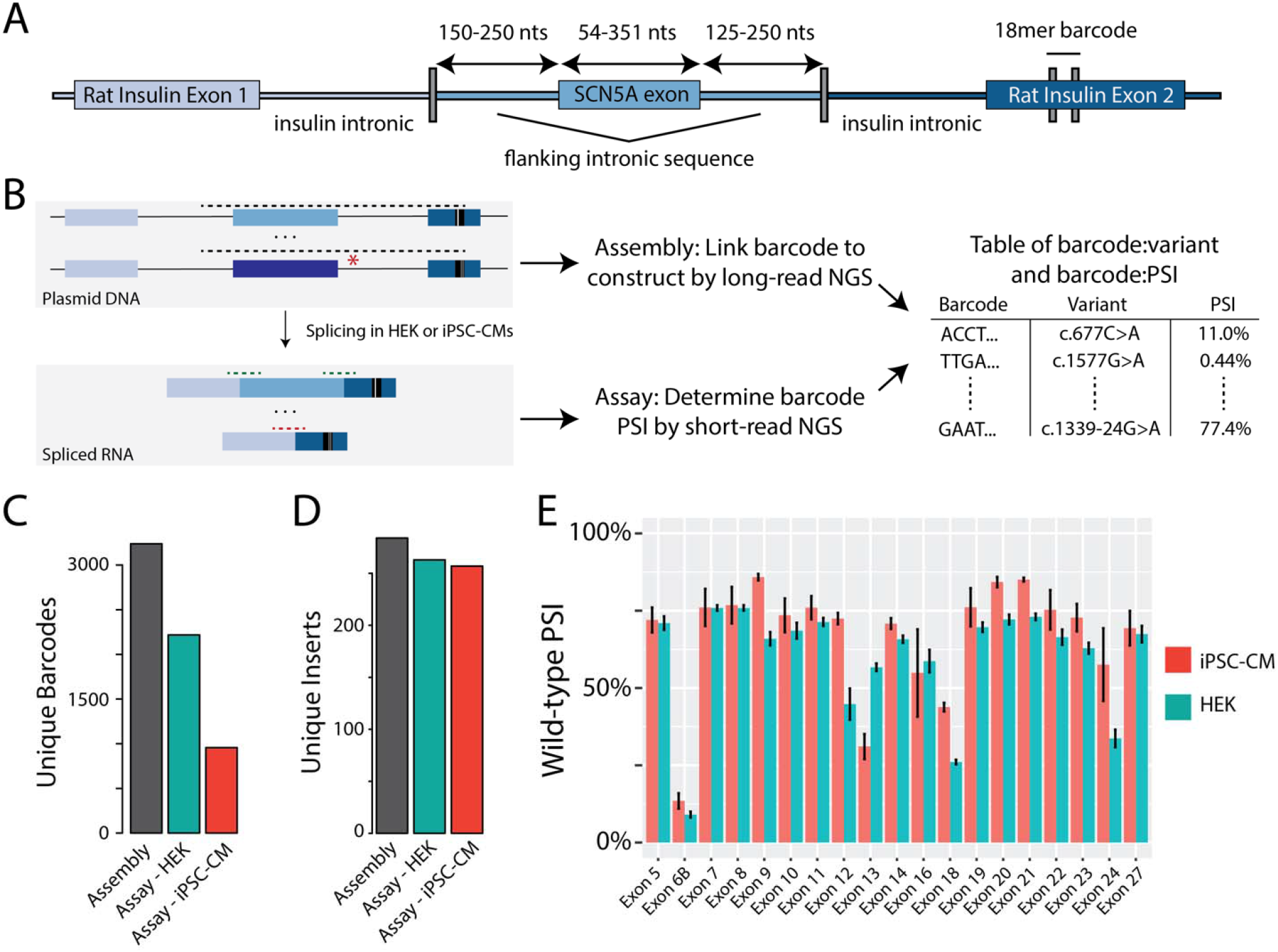
ParSE-seq assay in HEK cells and iPSC-CMs. A) Detailed schematic of the minigene plasmid. An 18-mer barcode inserts into downstream rat insulin exon 2. B) Overview schematic of assembly and assay steps, and subsequent integration. Dashed lines represent amplicons for long- and short-read sequencing. C) Barcode counts for the assembly, and recovered barcodes present across three replicates in HEK and iPSC-CM assays. D) Unique WT or variant inserts covered by barcodes in panel C. E) PSI for all WT exons in iPSC-CMs and HEK cells. Data is averaged across three replicates and error bars represent the standard error of the mean.

We transfected HEK cells and iPSC-CM cells each in triplicate with the barcoded library, isolated RNA, performed RT-PCR, and used targeted RNA-seq to quantify the impact of variants on minigene splicing. After quality control (see methods), we recovered splicing data in HEK cells for 2,218 barcodes associated with 263 unique inserts (244 variants and 19 WT exons, Figure 2C and 2D; raw data in Supplemental Table IV). Although the transfection efficiency in iPSC-CMs was low (∼2% of cells were GFP+), we nonetheless recovered splicing data for a total of 927 unique barcodes associated with 243 unique inserts (224 variants and 19 WT exons; Figure 2C and 2D; all PSI raw data in Supplemental Table V). PSI values between replicates were highly correlated (Supplemental Figures IV and V). We calculated a PSI for each WT exon in HEK and iPSC-CMs, and observed slightly higher PSIs in iPSC-CMs than in HEK cells (Figure 2E).

For each variant we calculated ΔPSI_norm, the normalized change in PSI of the variant compared to the PSI of the corresponding WT exon. Figures 3A-3C show the location and ΔPSI_norm of all variants quantified in the HEK and iPSC-CM assays. In Figure 3D, we show the continuous distribution of variant ΔPSI_norm among all studied variants in iPSC-CMs, which ranges from severe abrogation of splicing, to wildtype-like splicing, to enhanced PSI for a few variants (Figure 3C). Transfection of the barcoded library revealed excellent correlation of ΔPSI_norm scores between HEK cells and iPSC-CMs (*R*^*2*^ = 0.84, p = 6.4 * 10^−95^; Figure 3E). We considered variants with a false discovery rate (FDR) < 0.1 and a ΔPSI_norm < -50% to be ‘abnormal’, and variants to be ‘normal’ with an FDR > 0.1 and change in ΔPSI_norm > -20%; all other variants were considered indeterminate in our assay (Figure 3F). Additional thresholds for normal or abnormal ΔPSI based on 2 standard deviation differences of B/LB control variant normalized ΔPSI values are shown in Supplemental Figure VI, suggesting a more lenient PSI threshold of -0.36 for abnormal variants^26^. 105/94/45 variants had normal/abnormal/indeterminate splicing in the HEK293 assay, and 104/78/42 variants had normal/abnormal/indeterminate splicing in the iPSC-CM assay. Of the 244 and 224 variants studied in HEK and iPSC-CMs, 207 were present in both cell types. We observed no discordance in variant splicing results between the two cell types (variants with a normal splicing result in one cell type and an abnormal splicing result in the other cell type; Supplemental Figure VII). Due to the high concordance between assay results in the two cell types, we focused on the more physiologically-relevant iPSC-CM results for the rest of the analyses. We observed variable effects on splicing by variant class in Figure 3G. Among non-indeterminant variants, we observed splicing disruptions among 24/25 2-bp canonical splice sites variants, 20/48 missense variants, 18/57 synonymous variants, 5/6 stop-gain, and 11/45 non-canonical intronic variants.

**Figure 3.**
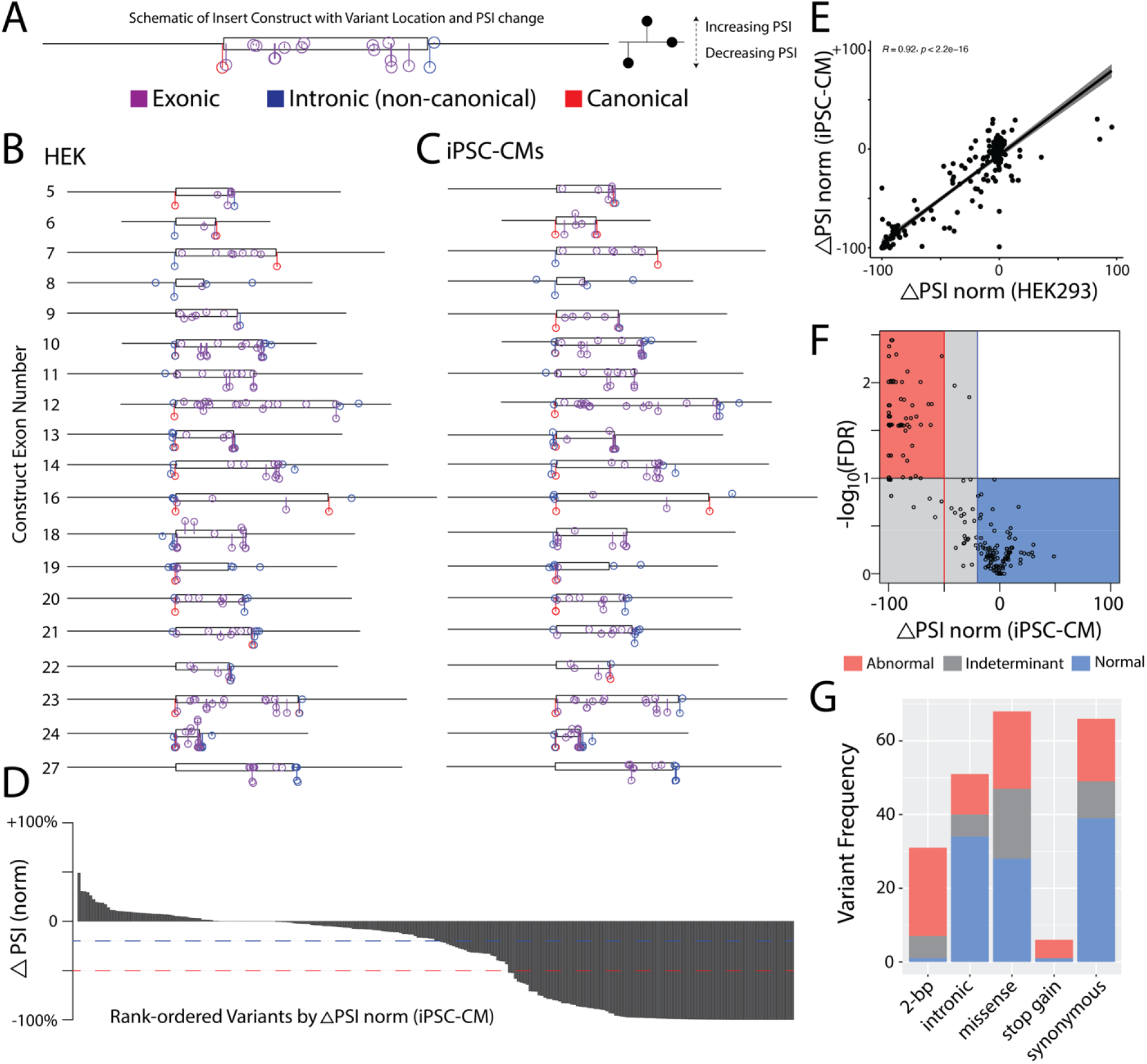
ParSE-seq results for a library of *SCN5A* variants. A) Example of variants superimposed along construct design – iPSC-CM construct 23 is shown. Y-axis corresponds to change in ΔPSI_norm, and X-axis position along the exonic (box) and intronic (line) segments of the synthetic insert. B) Distribution of ParSE-seq investigated variants in HEK cells. Change in y-axis corresponds to changes in ΔPSI_norm. C) Distribution of ParSE-seq investigated variants in iPSC-CMs. Change in y-axis corresponds to changes in ΔPSI_norm. D) Waterfall plot of ΔPSI_norm by variant. Red dashed line corresponds to -50% normalized ΔPSI, and blue to -20% normalized ΔPSI. E) Correlation of normalized ΔPSI between HEK and iPSC-CMs (n=207). F) Volcano plot of normalized ΔPSI and -log_10_(FDR). Each dot represents a variant studied in iPSC-CMs. G) Barplot of ParSE-seq variant outcomes by variant mutation type in iPSC-CMs.

### Functional Studies and SpliceAI

We next examined the concordance between our experimental splicing scores and the *in silico* predictor SpliceAI^5^. In Figure 4A, we show the distributions of the aggregate SpliceAI scores by ClinVar classification. Variants with higher SpliceAI scores were more likely be classified as P/LP compared to B/LB variants, with an intermediate distribution for VUS and CI variants. In Figure 4B, we plot ΔPSI_norm scores against aggregate SpliceAI scores. We observed a high correlation between SpliceAI scores and negative changes in PSI (*R*^*2*^ = 0.69, p = 3.9 * 10^−47^). Similar results were seen when canonical 2-bp splice variants were excluded (*R*^*2*^ = 0.56, p =4.6 * 10^−18^; Supplemental Figure VIII). Assessing the dichotomous outcome of normal or abnormal splicing, we observed high sensitivity and specificity of SpliceAI predictions, reaching an area under the curve (AUC) of 0.956 across the full library (Figure 4C). Notably, there was high concordance of SpliceAI predictions and ParSE-seq scores for the subclasses of exonic variants (AUC=0.947) or intronic variants outside of the canonical splice sites (AUC=0.989). Across quintiles of SpliceAI scores, we observed different fractions of normal/abnormal/indeterminant variants, with the highest fraction of indeterminant variants in the intermediate 3 quintiles (14/51) indeterminant variants; Figure 4D). To further explore the robustness of SpliceAI, we evaluated its ability to predict splice-altering exonic variants. Overall, 41 variants with pre-computed SpliceAI scores >0.80 had normal or abnormal splicing in the iPSC-CM assay, including two ClinVar VUS’s (c.1338G>A/p.Glu446Glu, ClinVar: 451631 and c.4297G>T/p.Gly1433Trp, ClinVar: 519412). ParSE-seq outcomes of these 41 variants are shown in Figure 4E, suggesting a high positive predictive value of SpliceAI-prioritized variants.

**Figure 4.**
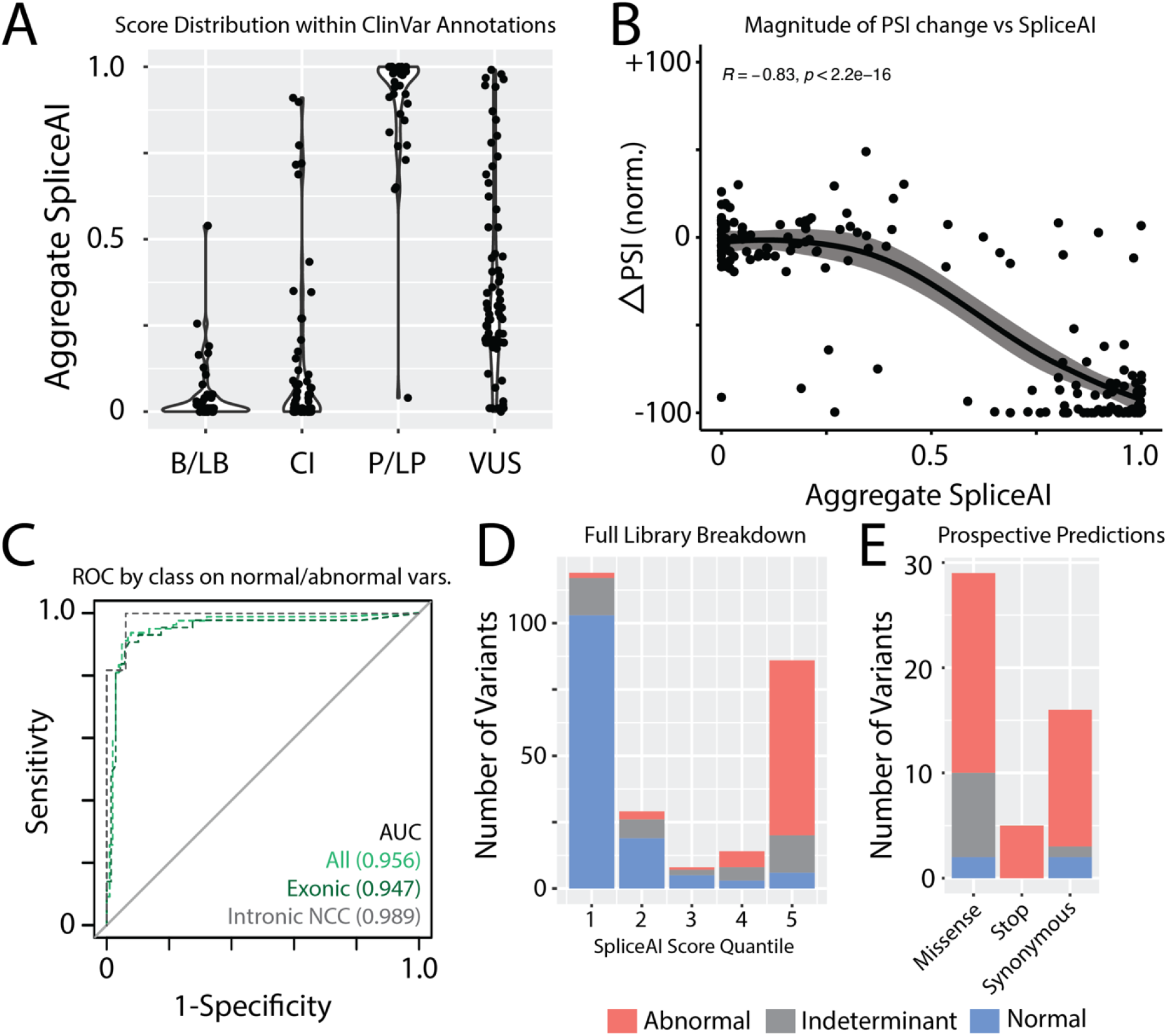
Comparison of experimental data and *in silico* SpliceAI scores. A) Aggregate SpliceAI scores for each ClinVar variant class. B) Correlation of normalized ΔPSI against aggregate SpliceAI scores. Confidence interval fit using Loess. C) Receiver operating characteristic curves for SpliceAI applied to normal and abnormal variants. Indeterminant variants are not included. AUC=area under the curve, NCC – non-canonical splice sites. D) Distribution of variant effect in ParSE-seq stratified by SpliceAI score quantiles. E) Results of prospectively identified exonic variants by SpliceAI score >0.8 stratified by mutation type and ParSE-seq outcome.

### ACMG Assay Calibration

We tested ClinVar B/LB and P/LP control variants to provide a calibrated strength of evidence for the assay. We followed the ClinGen Sequence Variant Interpretation Working Group scheme for converting control assay results into likelihood ratios of pathogenicity, termed “OddsPath”^24^. A total of 47 ClinVar B/LB and P/LP control variants were recovered from our splicing experiment in iPSC-CMs with normal or abnormal splicing results. We observed a near-perfect concordance of ClinVar classifications and assay outcomes in iPSC-CMs: 24 of 25 P/LP variants had abnormal splicing and 21 of 22 B/LB variants had normal splicing (Figure 5A; detailed math in Supplemental File I). Applying these values, the OddsPath_Pathogenic_ (strength of evidence of abnormal assay results) was 21.1 and the OddsPath_Benign_ (strength of evidence of normal assay results) was 0.042. These values correspond to strong application of abnormal functional evidence towards pathogenicity (PS3 strong; OddsPath > 18.7) and strong application of normal functional evidence towards a benign classification (BS3 strong; OddsPath <0.053). The HEK cell assay performed similarly well with 30/31 concordant P/LP variants and 27/27 concordant B/LB variants, yielding a OddsPath_Pathogenic_ of 26.13 and an OddsPath_Benign_ of 0.032 (PS3 strong and BS3 strong; Figure 5B). The discrepant B/LB control in iPSC-CMs was an indeterminant variant in the HEK cell assay, with a ΔPSI_norm of 0.36 and FDR of 0.001 in HEK293 cells.

**Figure 5.**
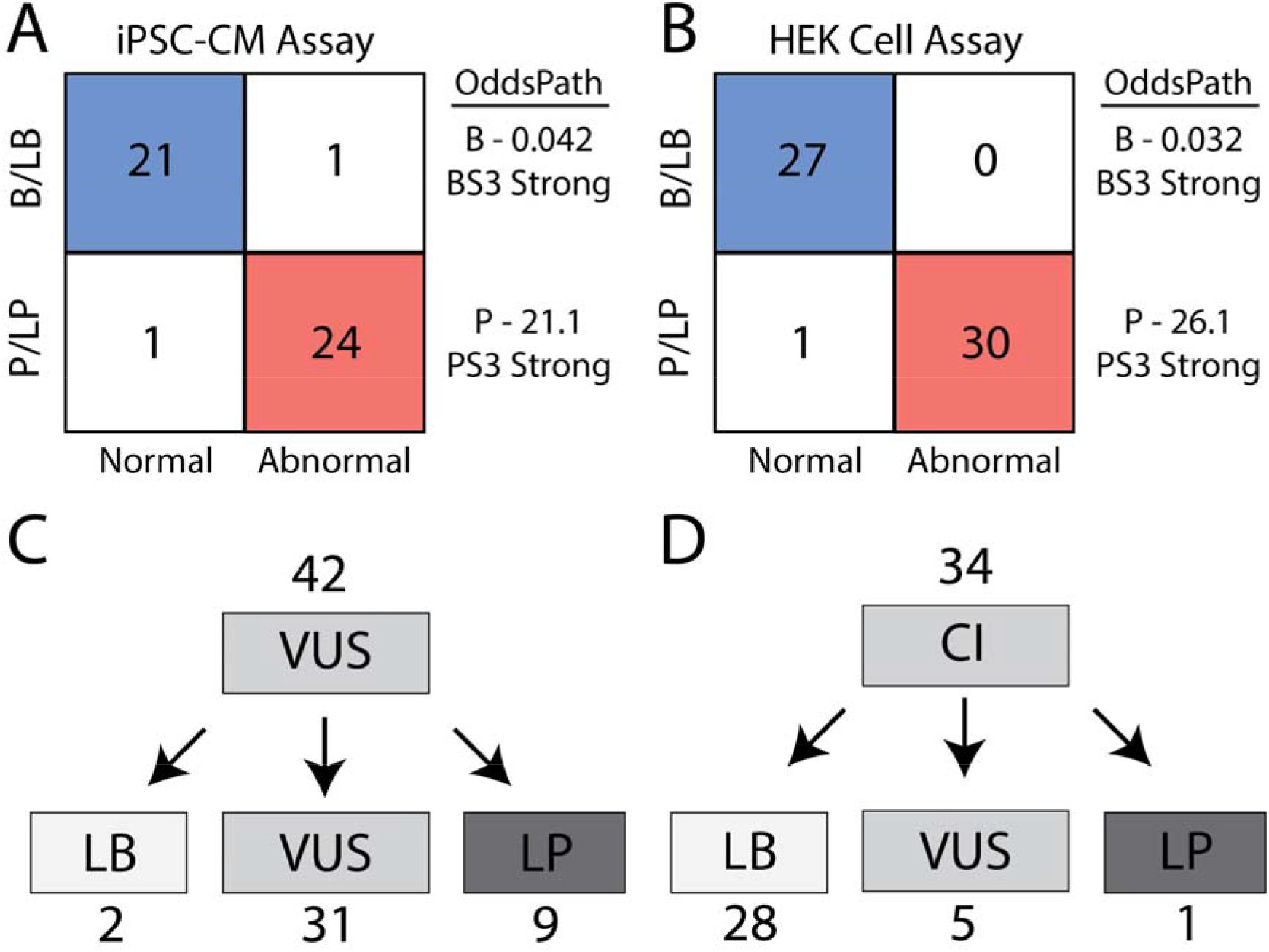
Assay calibration and evidence-based variant classification. A) ParSE-seq results for B/LB and P/LP controls in iPSC-CMs. As described in the methods, we obtain 2 OddsPath values to implement BS3 and PS3, both at a strong level of evidence. B) ParSE-seq results for B/LB and P/LP controls in HEK cells. As described in the methods, we obtain 2 OddsPath values to implement BS3 and PS3, both at a strong level of evidence. C) VUS were reclassified using functional data from ParSE-seq at the strong level of evidence. BS3 evidence was applied exclusively to synonymous and intronic VUS. PS3 evidence is applied to all VUS. D) Classifications of Conflicting Interpretation variants using functional evidence. BS3/PS3 applied as for VUS in Panel C.

### Variant Reclassification

After identifying a calibrated strength of evidence to implement functional data (PS3 strong and BS3 strong), we sought to provide updated clinical interpretations for 42 VUS and 34 CI variants for which we obtained normal or abnormal splicing results in the iPSC-CM assay. We identified 8 VUS that were abnormal in our splicing assay (5 exonic, 3 intronic). Applying these functional data and *in silico* predictions from SpliceAI, we reclassified 9 abnormally splicing variants to LP, and 2 normally splicing variant to LB (Figure 5C; all ACMG criteria in Supplemental Table VI). Since a normal splicing result in the ParSE-seq assay does not address possible dysfunction of the translated protein product for missense variants, we limited application of benign functional data to synonymous and intronic variants. Variants with annotations of “Conflicting Interpretations” are a growing fraction of variants in ClinVar^60,61^. We anticipated that functional studies could provide new data to adjudicate variants with conflicting interpretations, especially for synonymous and intronic variants. Applying the ParSE-seq results from iPSC-CMs, we classified 28 of 34 CI variants as LB (Figure 5D). We classified 1 CI variant as LP due to its splice-altering effect.

### Cryptic splicing effects of missense variants

Missense variants that disrupt gene function and lead to disease are usually presumed to disrupt protein function. Functional assays of missense variants are often performed using cDNA, which obscures variant effects on splicing.^62^ We hypothesized that some *SCN5A* missense variants may cause Mendelian disorders such as BrS through an aberrant splicing mechanism rather than disruption of protein function (Figure 6A). In iPSC-CMs, we recovered determinate data for 48 missense variants; 28 were listed as VUS in ClinVar, and 20 were identified prospectively with high SpliceAI scores (>0.8). Of these, we identified 18 splice-altering missense variants, created by 20 unique splice-altering single nucleotide variants. Figure 6B shows the spatial distribution of the missense variants across SCN5A. These variants were distributed throughout the protein, but often clustered in hotspots near exon boundaries. We studied 2 missense variants in further detail: c.4220C>G/p.A1407G (ClinVar: 846026) and c.3392C>T/p.T1131I (ClinVar: 67795). These variants were both associated with Brugada Syndrome in ClinVar and disrupted splicing in the ParSE-seq assay (Figure 6C). We performed automated patch clamping experiments on these two variants using HEK293 cells stably expressing *SCN5A* cDNA. The two missense VUS, both had near-normal electrophysiologic function in the cDNA-based electrophysiological assay—normalized peak current densities of 106.1±16.8% (n=19) and 85.4±9.5% (n=41) of WT, respectively (Figure 6D). We next compared our experimental splice-altering missense variant results with two *in silico* predictors specifically for missense variants – a Bayesian structural penetrance prediction model^45^ and REVEL^44^. First, a continuous method that places high weights on protein structure was moderately sensitive to detect pathogenic signal among splice-altering missense variation 12/20 (Figure 6E). In contrast, the ensemble predictor REVEL would classify 17/20 splice-altering missense variants as pathogenic at a conservative threshold (Figure 6E). Thus, ParSE-seq can help identify a class of missense, splice-altering variants for which cDNA-based assays of protein function yield incorrect conclusions about variant pathogenicity.

**Figure 6.**
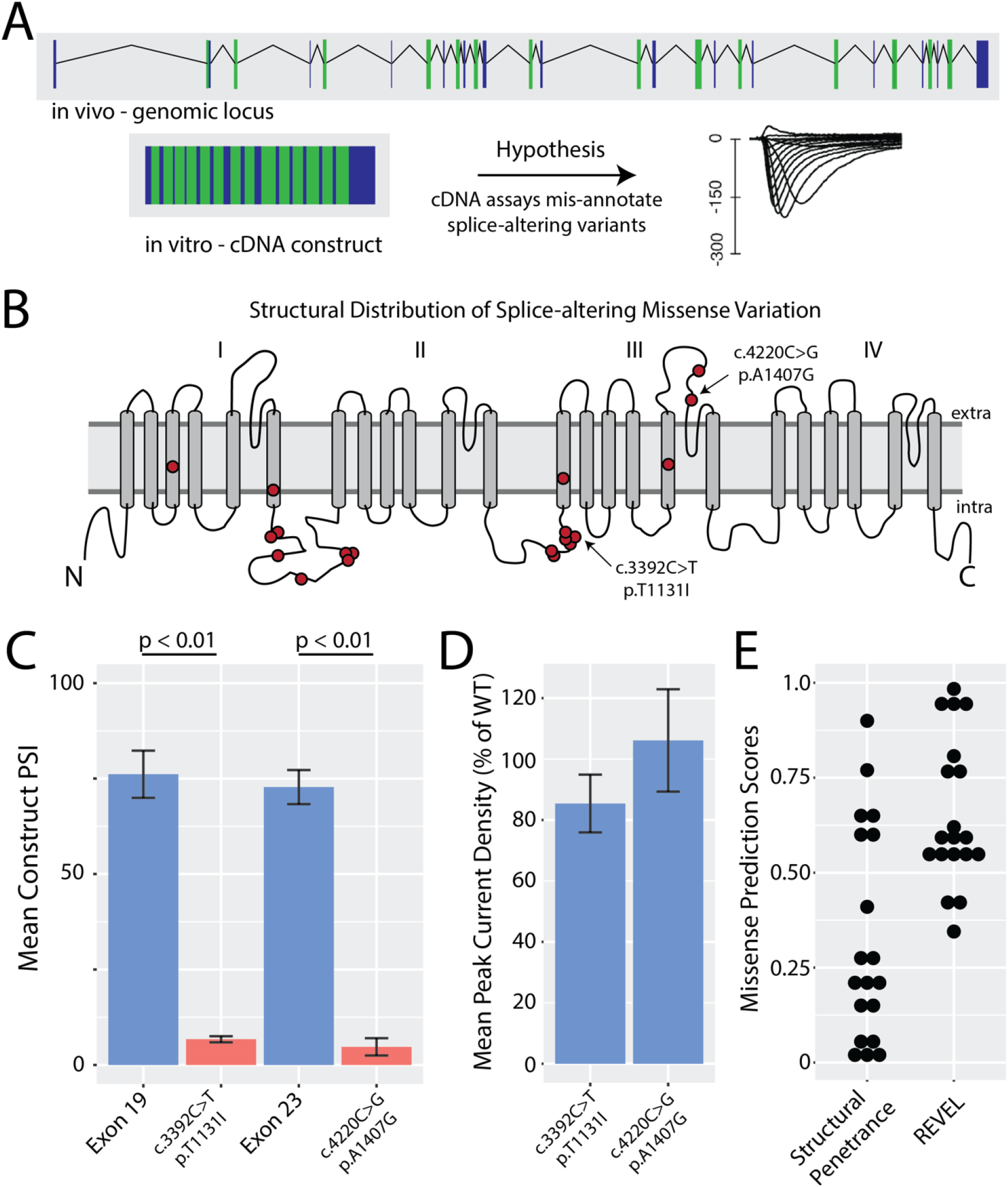
Investigation of missense variants with ParSE-seq and automated patch clamp studies. A) Schematic of genomic locus vs cDNA-based plasmid used in many *SCN5A* functional assays. These assays do not account for splice-altering variant effects. B) Structural distribution of splice-altering missense variants in *SCN5A* revealed by ParSE-seq. C) ParSE-seq results for two splice-altering variants compared with their corresponding wildtype exons (n=3 per group; error bars correspond to standard error of the mean, p < 0.01 [2-sided t-test]). D) Plots of missense variant peak current densities normalized to WT. Variants were studied with the SyncroPatch 384 PE automated patch clamp system compared to WT. 19-41 cells studied per variant. Error bars correspond to standard error of the mean. E) Scores of *in silico* predictors from Bayesian structural penetrance strategy and REVEL. Bayes score >0.2 indicates higher risk of pathogenicity. REVEL scores >0.5 are considered pathogenic.

### Recapitulating Splice Effects at the Endogenous Locus

Modeling cardiac disease has been greatly advanced by developments in genome editing and iPSC-CM differentiation techniques^63^. We further studied an *SCN5A* intronic variant, c.1891-5C>G (ClinVar: 201463), which disrupted splicing in the ParSE-seq minigene assay (ΔPSI_norm = -79%, FDR=0.046). We used CRISPR-Cas9 to introduce this variant as a heterozygous edit into a healthy control iPSC line (Figure 7A). This variant was predicted by SpliceAI to use the competing ‘AG’ motif formed by this single nucleotide variant (Figure 7B+7C). The hemizygous ParSE-seq overexpression assay showed significant quantities of a predicted 4-nucleotide indel among the variant construct that is predicted to cause a frameshift (Figure 7D). To avoid confounding by nonsense-mediated decay (NMD) in the endogenous locus of heterozygous iPSC-CMs^64,65^, we treated both isogenic lines with the NMD inhibitor cycloheximide (CHX). We performed RNA-seq and were able to observe a small fraction of the 4-nucleotide retention event in the heterozygous line treated with CHX, but not in control lines or DMSO-treated heterozygous cells, consistent with NMD of the aberrant transcript^54,64^ (Figure 7E). To test these effects on protein-level properties, isogenic pairs of iPSC-CMs were studied by patch clamping to measure sodium channel current (Figures 7F+7G). We observed a large decrease in peak current in the edited lines, confirming the loss-of-function phenotype typical for *SCN5A*-linked BrS^66,67^ (Figure 7H).

**Figure 7.**
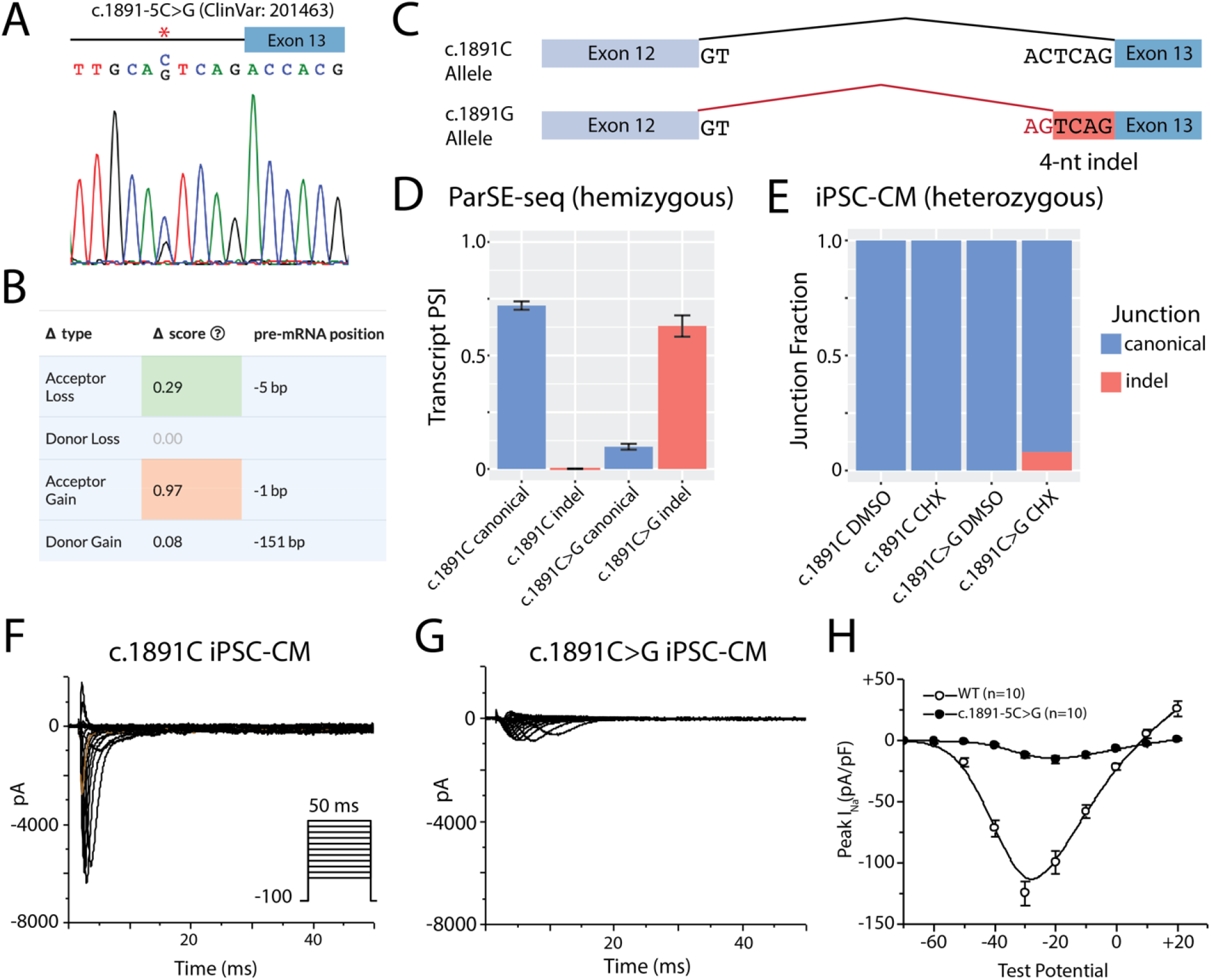
Intronic variant at the endogenous *SCN5A* locus disrupts *SCN5A* RNA splicing and protein-level sodium current. A) The c.1891-5C>G variant was introduced as a heterozygous edit with CRISPR-Cas9 into a control iPSC line. Both control and edited lines underwent differentiation to iPSC-CMs. B) Variant level SpliceAI predictions of the c.1891C>G variant. New motif is predicted to abolish canonical splice site and introduce a novel splice site at the *de novo* AG motif. C) Schematic depiction of splicing outcomes for the WT allele and variant allele. The variant allele is predicted to use a cryptic splice site and retain 4 nucleotides of intronic sequence as a frame-shifting indel. D) Hemizygous ParSE-seq results of the PSI for the WT splicing product vs the specifically predicted 4-bp indel created by the competing AG motif in the WT and variant constructs. E) Fraction of canonical vs ParSE-seq predicted indel in WT lines and heterozygous edited lines. Blue indicates canonical splicing, whereas red indicated the 4-nucleotide pseudoexon. F) Sodium current traces of the healthy control line. Voltage protocol provided in inset. G) Sodium current traces of the heterozygous c.1891C>G line. Same voltage protocol as in F. H) Quantification of peak current at -30 mV of the isogenic lines. p < 0.01 (2-sided t-test).

## Discussion

### ParSE-seq Summary

In this study, we developed a high-throughput method to assess the splicing consequences of hundreds of variants (ParSE-seq). We implemented barcoding of a pool of minigene plasmids to enable multiplexed splicing readouts using high-throughput sequencing and applied the method to study variants in the cardiac sodium channel gene *SCN5A*. In iPSC-CMs, we quantify variant effects on splicing for 224 variants, and detected 78 variants with abnormal splicing. We observed concordance of splicing results for 45/47 B/LB and P/LP variants, and we determined that our assay could be applied at the strong level in the ACMG classification scheme (BS3 and PS3) when assuming ClinVar as ground truth. Leveraging these calibrated strengths of evidence, we classified *SCN5A* VUS and CI variants. We determined that the *in silico* tool SpliceAI has excellent, but not perfect, concordance with experimentally measured splicing effects. We also demonstrated examples of missense variants that had normal electrophysiology using conventional heterologous expression cDNA-based approaches but disrupt splicing. Lastly, we showed that our ParSE-seq results predict aberrant splicing in a disease relevant iPSC-CM model, with consequences at both the RNA and protein levels. We envision that ParSE-seq will be applicable to many disease genes and will be accessible using openly available computational pipelines and democratized gene synthesis available to the community.

### Incorporating Splicing Assays into the Genetic Testing Pipeline

A recent survey of clinical patient variant data highlighted the need for accurate splicing assays to improve variant classification and support the implementation of personalized medicine^2^. Our assay identified 43 exonic variants and 11 intronic variants outside canonical splice sites which disrupt splicing in iPSC-CMs. This data is of immediate clinical relevance to individuals who are heterozygous for these variants, and they should be screened for BrS and cardiac conduction defects seen with *SCN5A* loss-of-function variants. In addition, we identified 30 VUS or CI variants that could be classified as LB after incorporating our splicing data, which reduces clinical uncertainty for these variant carriers^2^. Recently, standardization of techniques and harmonization of classifications have improved the clinical integration of splicing results from primary or peripheral tissue^9^. We anticipate that lab-based methods like ParSE-seq and clinical studies of peripheral tissue will be complementary. *In vitro* functional data will be especially useful when tissue is not available, when there are no other heterozygotes to segregate a phenotype, or expression of the relevant transcript is not sufficiently high in the clinically accessible tissue. Clinical potential will be further improved by the integration of functional assays and AI-based *in silico* tools for splicing predictions^5,68^.

### Comparison to Other High-throughput Splicing Assays

ParSE-seq builds on pioneering work in the field of high-throughput splicing assays. For example, multiplexed splicing readouts were recently applied to *LMNA, MYBPC3*, and *TTN* variants using minigene-based large oligo constructs and barcoding^20,21^. Although some previous studies have examined splice outcomes for thousands of multiplexed variants, these experiments are typically limited to only small exons with minimal adjacent intronic sequence due to the size limits of chip-based oligonucleotide synthesis^6,18,19,69^. This approach often necessitates the inclusion of only small adjacent intronic regions and precludes studying large exons. ParSE-seq builds on these approaches by considering a more complete set of aberrant splicing patterns through the triplet exon cassette design, as well as enabling the testing of much larger clinically relevant exons feasible with commercially-available clonal gene synthesis technology. For most tested exons we included 250 bp of intronic sequence on each side of the target exon. Inclusion of larger segments of flanking introns has been shown to increase assay validity due to more complete capture of *cis*-regulatory elements^18^. A major advantage of this framework is the standardization of library preparation, wherein a library can quickly be assembled using genomic coordinates of variants of interest. In addition, previous high-throughput splicing methods have not been calibrated using known B/LB and P/LP standards for clinical variant classification in the ACMG scheme. Further, we perform the assay in a disease-relevant cell type (iPSC-CMs).

ParSE-seq may also complement multiplexed assays of variant effect (MAVEs), proactive high-throughput assays of variant function^70^. For example, MAVEs subject libraries of many variants at each position of a protein to a fitness assay, which allows prospective functional evaluation of a variant even before it is clinically observed. This approach holds great promise for reducing the VUS problem^71^. One limitation is that many previous approaches have used cDNA-based libraries. Here we show the limits of these approaches for certain missense variants, which would produce disparate outcomes depending on the assay. In some previous cDNA-based deep mutational scans, investigators have excluded their functional results for variants with SpliceAI scores above a certain threshold^72^, or assumed deleterious splicing based on prediction-alone when applying results to clinical cohorts^27^. We now show the feasibility of obtaining prospective splicing results of many exonic variants prioritized by an *in silico* predictor. Comprehensive splicing data for predicted splice-altering variants in cardiac channelopathy genes could complement cDNA-based MAVEs in progress for these genes.^73,74^

### Future Directions

ParSE-seq can rapidly assess hundreds of candidate splice-altering variants. Given the plethora of VUS and CI variants that may disrupt splicing^2^, this method may help classify large sets of variants in Mendelian disease-associated genes that act through a loss of function mechanism.

### Limitations

Although we validate one splice-altering variant by CRISPR editing of the endogenous locus, most variants were tested only in minigene assays. There may be examples where the ParSE-seq minigene assay does not fully capture all nuances of biology at the endogenous locus. The first and last exon, exons using non-canonical 2-bp splice sites, and exons that were difficult to synthesize due to high GC content were not included in the library.

### Conclusions

We describe ParSE-seq, an ACMG-calibrated high-throughput splicing assay. We anticipate that ParSE-seq will be a useful method for to rapidly assessing variant splicing effects. Given the plethora of variants that may act through disrupting splicing, this method may quickly characterize the splicing effects of variants in disease-associated genes.

## Data Availability

All data produced in the present study are available upon reasonable request to the authors.

https://github.com/GlazerLab/ParSE-seq

## Description of Supplemental Data

We have included all referenced supplemental figures and tables (Supplemental Figures I-VIII, Supplemental Tables I-VI, and Supplemental File I) in the accompanying Supplement.

## Ethics Statement

Relevant human subjects work was approved by the Vanderbilt University Medical Center IRB #9047.

## Acknowledgements

This research was funded by American Heart Association fellowship 907581 (MJO), and by the National Institutes of Health: 1F30HL163923-01 (MJO), T32GM007347 (MJO), R00 HG010904 (AMG), R35 GM150465 (AMG), R01 HL149826 (DMR), and R01 HL164675 (DMR). Flow cytometry experiments were performed in the Vanderbilt Flow Cytometry Shared Resource. The Vanderbilt Flow Cytometry Shared Resource is supported by the Vanderbilt Ingram Cancer Center (P30 CA68485) and the Vanderbilt Digestive Disease Research Center (DK058404). The Nanion SyncroPatch 384PE is housed and managed within the Vanderbilt High-Throughput Screening Core Facility, an institutionally supported core, and was funded by NIH Shared Instrumentation Grant 1S10OD025281. The HTS Core receives support from the Vanderbilt Institute of Chemical Biology and the Vanderbilt Ingram Cancer Center (P30 CA68485). We thank Maryland Genomics at the Institute for Genome Sciences, University of Maryland School of Medicine for performing PacBio SMRT sequencing. We thank Kenneth Matreyek and Doug Fowler for the HEK293 landing pad cells. We thank Yuko Wada and Lorena Harvey for assistance with genome editing.

## Data and Code Availability

Code used to analyze raw data and generate figures and tables are available at the Glazer Lab GitHub site. All DNA sequencing data will be made available at the NCBI Sequence Read Archive at publication. Variant classifications and functional data will be uploaded to ClinVar upon publication.

