## Supplemental Material for "ParSE-seq: A Calibrated Multiplexed Assay to Facilitate the Clinical Classification of Putative Splice-altering Variants"

Table of Contents

**Supplemental Methods.** Description of NGS datasets used in this study.

**Supplemental Figure I.** Schematic of minigene vector design, primers, and restriction sites.

**Supplemental Figure II.** Schematic of assay and assembly computational workflows.

**Supplemental Figure III.** Library barcode frequency by Illumina and PacBio sequencing methods.

**Supplemental Figure IV.** PSI correlations among HEK replicates.

**Supplemental Figure V.** PSI correlations among iPSC-CM replicates.

**Supplemental Figure VI.** Derivation of ΔPSI norm. thresholds of benign variants for calibration.

**Supplemental Figure VII**. Variant interpretations between HEK and iPSC-CM assays.

**Supplemental Figure VIII.** ParSE-seq vs SpliceAI w/o 2-bp slice variants.

**Supplemental Table I.** Primers used in this study.

**Supplemental Table II.** Construct exon and flanking intron lengths.

**Supplemental Table III*.** Variant characteristics – genomic position, gnomAD allele frequency, ClinVar classification, and SpliceAI prediction.

**Supplemental Table IIV*.** ParSE-seq HEK cell data for all studied variants.

**Supplemental Table V*.** ParSE-seq iPSC-CM cell data for all studied variants.

**Supplemental Table VI*.** ACMG criteria for variant interpretation.

**Supplemental File I***. Excel calculation of OddsPath.

***Large table included in separate excel attachment**

**Supplemental Methods**

*Description of Next Generation Sequencing datasets used in this study.* The following are descriptions of the NGS datasets used in the ParSE-seq manuscript. All DNA sequencing data will be made available at the NCBI Sequence Read Archive at publication.

**MO-9293** – PacBio long-read sequencing dataset for assembly.

**MO-9402** – Illumina short-read sequencing of barcode frequency from barcoded plasmid pool. Used to compare barcode abundance based on long-read sequencing and short read sequencing (Supplemental Figure II).

**MO-9419** – Illumina short-read sequencing of assay performed in triplicate in iPSC-CMs.

**MO-9489** – Illumina short-read sequencing of assay performed in triplicate in HEK cells.

**MO-9518** – Illumina RNA-seq data for wildtype iPSC-CMs treated with CHX and DMSO.

**MO-9872** – Illumina RNA-seq data for heterozygous c.1891-5G>C iPSC-CMs treated with CHX and DMSO.

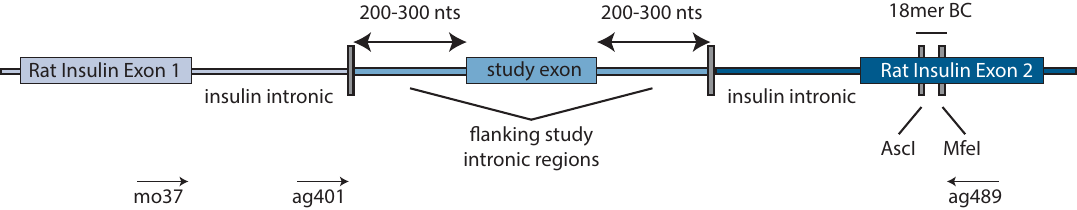

**Supplemental Figure I. Schematic of minigene vector design, primers, and restriction sites**. Study region was synthesized and cloned into pAG424 by Twist Biosciences. An 18-mer barcode is positioned in the downstream pET01 vector. Primers are designed to allow for assay completion by short-read NGS (mo37 and ag401) and assembly by long-read NGS (mo37 and ag489).

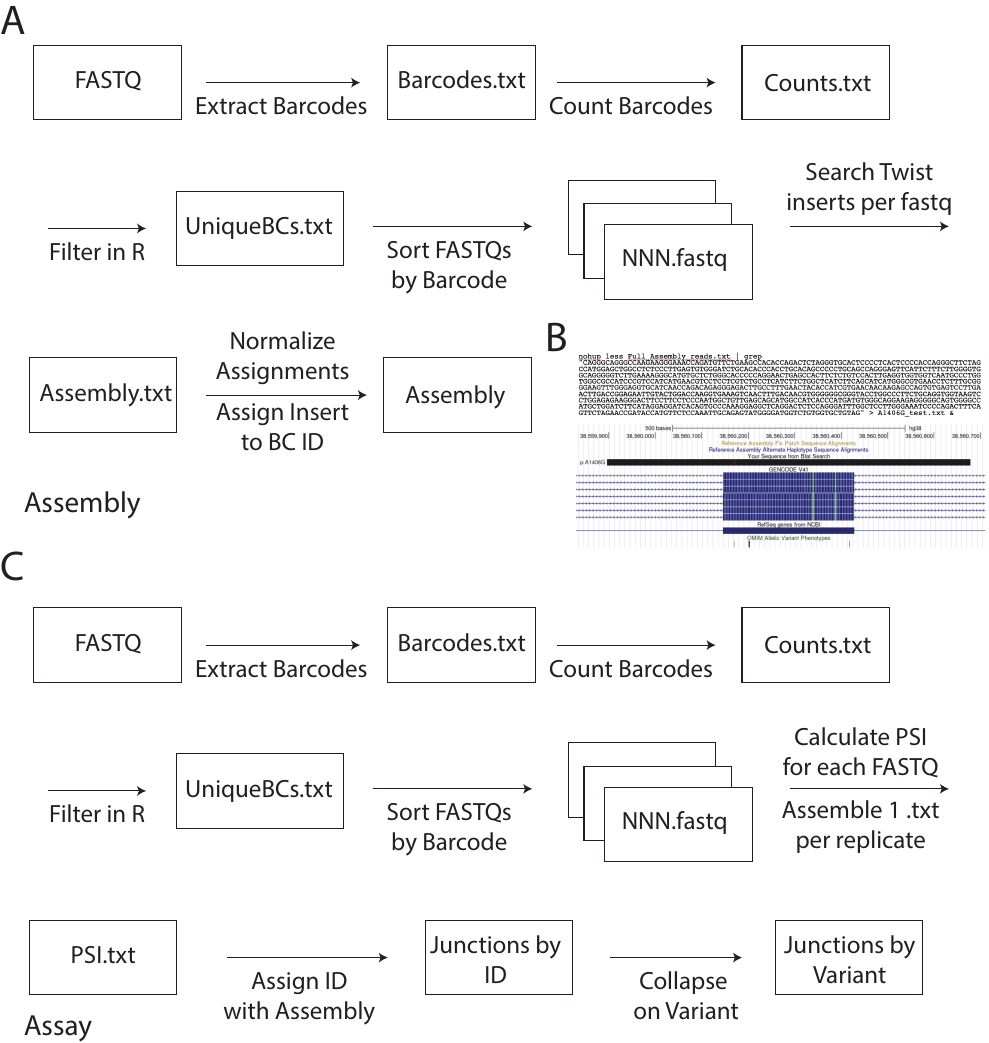

**Supplemental Figure II. Schematic of assay and assembly computational workflows.**

**A)** Assembly process from raw FASTQ to assembly cipher.

**B)** Genome Browser representation of searched Twist insert showing variant.

**C)** Assay process from raw FASTQ to processed reads per variant.

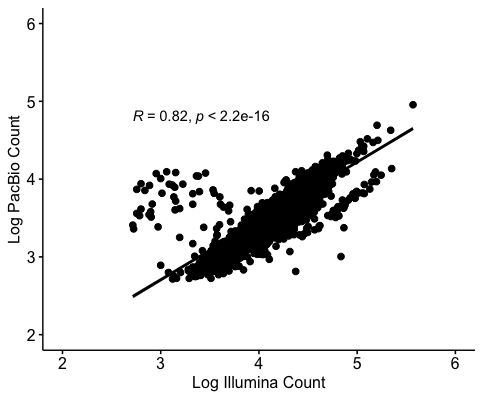

**Supplemental Figure III. Library barcode frequency by Illumina and PacBio sequencing methods**. Pearson correlation of barcode counts of pooled barcoded plasmids by Illumina sequencing and PacBio sequencing. Each dot represents one unique barcode.

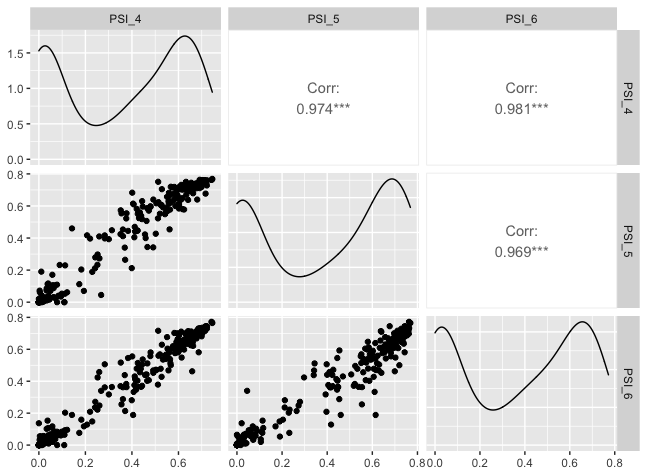

**Supplemental Figure IV.** **PSI correlations among HEK replicates.** Spearman correlations among ParSE-seq replicates in HEK cell assay experiments using ggpairs function in R.

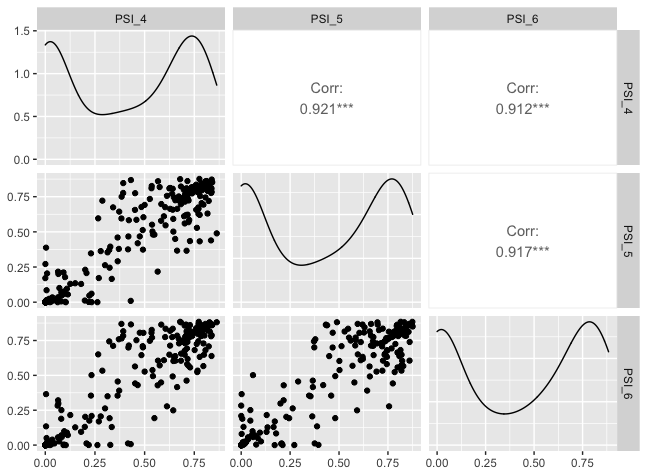

**Supplemental Figure V.** **PSI correlations among iPSC-CM replicates.** Spearman correlations among ParSE-seq replicates in iPSC-CMs assay experiments using ggpairs function in R.

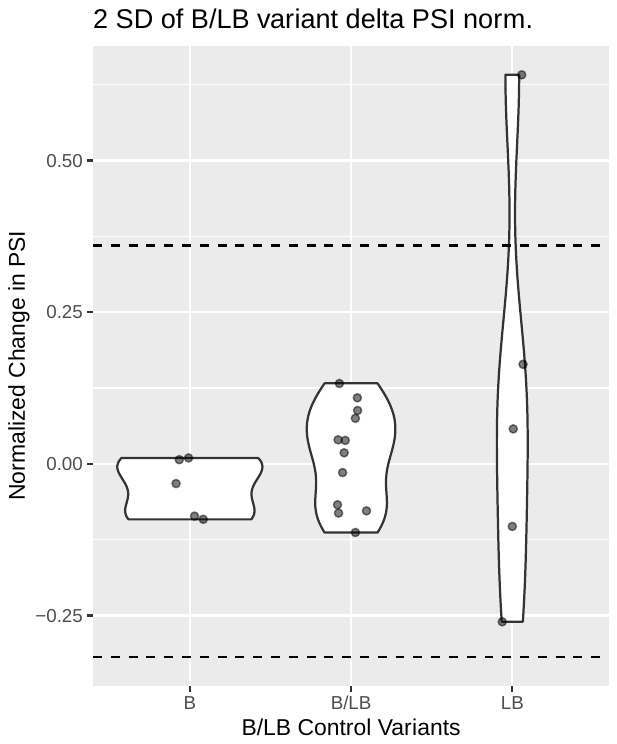

**Supplemental Figure VI. Derivation of ΔPSI norm. thresholds of benign variants for calibration.** Mean ΔPSI norm among controls is 0.02 with standard deviation of 0.169.

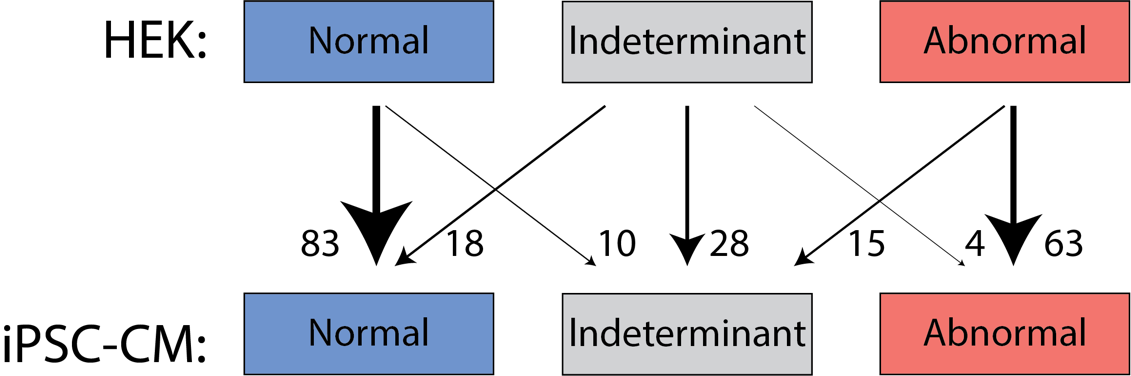

**Supplemental Figure VII.** Variant interpretations between HEK and iPSC-CM assays. There were no variants with a normal score in one cell type and an abnormal score in the other cell type.

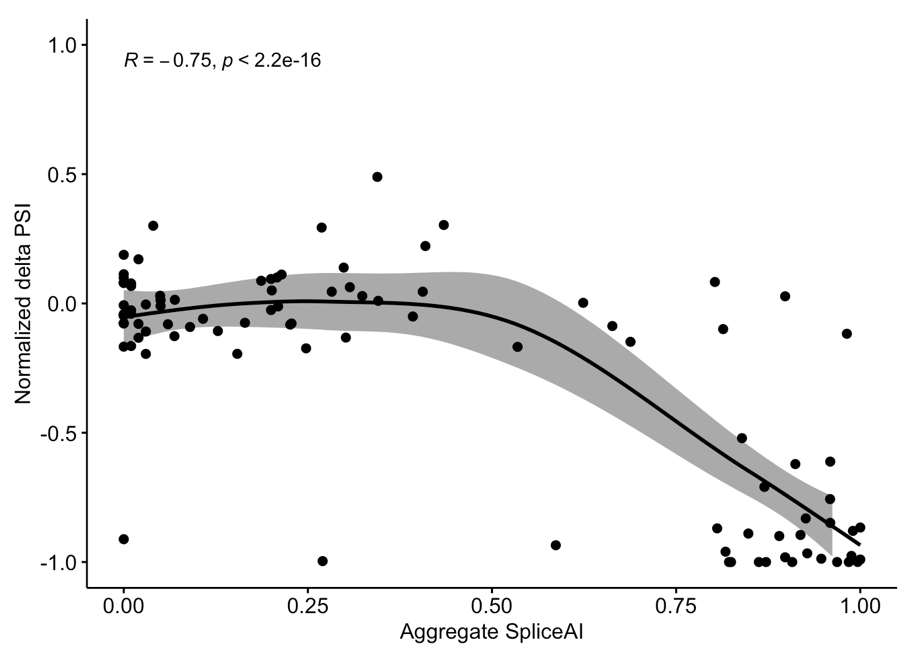

**Supplemental Figure VIII.** ParSE-seq and SpliceAI correlation by LOESS without inclusion of canonical 2-bp splice site-disrupting variants.

**Supplementary Table I.** Primers used in this study.

| **Primer** | **Sequence** | **Description** |
| --- | --- | --- |
| ag1799 | CAAGCAGAAGACGGCATACGAGATCGGTCCGATAGTGACTGGAGTTCAGACGTGTGCTCTTCCGATCTGCGTGGATTCTTCTACACACCC | Illumina I7 primer |
| ag1800 | CAAGCAGAAGACGGCATACGAGATGCCGGCGTATGTGACTGGAGTTCAGACGTGTGCTCTTCCGATCTGCGTGGATTCTTCTACACACCC | Illumina I7 primer |
| ag1801 | CAAGCAGAAGACGGCATACGAGATATTAATACGCGTGACTGGAGTTCAGACGTGTGCTCTTCCGATCTGCGTGGATTCTTCTACACACCC | Illumina I7 primer |
| ag1802 | CAAGCAGAAGACGGCATACGAGATGTTGTGTAACGTGACTGGAGTTCAGACGTGTGCTCTTCCGATCTGCGTGGATTCTTCTACACACCC | Illumina I7 primer |
| ag1803 | CAAGCAGAAGACGGCATACGAGATACCACACGGTGTGACTGGAGTTCAGACGTGTGCTCTTCCGATCTGCGTGGATTCTTCTACACACCC | Illumina I7 primer |
| ag1804 | CAAGCAGAAGACGGCATACGAGATTTGTAGTGTAGTGACTGGAGTTCAGACGTGTGCTCTTCCGATCTGCGTGGATTCTTCTACACACCC | Illumina I7 primer |
| ag1805 | CAAGCAGAAGACGGCATACGAGATCCACGACACGGTGACTGGAGTTCAGACGTGTGCTCTTCCGATCTGCGTGGATTCTTCTACACACCC | Illumina I7 primer |
| ag1806 | CAAGCAGAAGACGGCATACGAGATGGCAGTAGCAGTGACTGGAGTTCAGACGTGTGCTCTTCCGATCTGCGTGGATTCTTCTACACACCC | Illumina I7 primer |
| ag1807 | CAAGCAGAAGACGGCATACGAGATAATGACGATGGTGACTGGAGTTCAGACGTGTGCTCTTCCGATCTGCGTGGATTCTTCTACACACCC | Illumina I7 primer |
| ag1808 | CAAGCAGAAGACGGCATACGAGATTTGACCAATGGTGACTGGAGTTCAGACGTGTGCTCTTCCGATCTCTGGATTGTCCTATGTGTCTTGCTT | Illumina i7 primer |
| ag1809 | AATGATACGGCGACCACCGAGATCTACACGAGCGACGATACACTCTTTCCCTACACGACGCTCTTCCGATCTGGGCCACCTCCAGTGC | Illumina i5 primer |
| ag1646 | AATGATACGGCGACCACCGAGATCTACACCCGTATGTTCACACTCTTTCCCTACACGACGCTCTTCCGATCTGGGCCACCTCCAGTGC | Illumina I5 Primer |
| ag1647 | AATGATACGGCGACCACCGAGATCTACACAAGATACACGACACTCTTTCCCTACACGACGCTCTTCCGATCTGGGCCACCTCCAGTGC | Illumina I5 Primer |
| ag1648 | AATGATACGGCGACCACCGAGATCTACACGGAGCGTGTAACACTCTTTCCCTACACGACGCTCTTCCGATCTGGGCCACCTCCAGTGC | Illumina I5 Primer |
| ag1649 | AATGATACGGCGACCACCGAGATCTACACATTGATACTGACACTCTTTCCCTACACGACGCTCTTCCGATCTGGGCCACCTCCAGTGC | Illumina I5 Primer |
| ag1650 | AATGATACGGCGACCACCGAGATCTACACGCCAGCGTCAACACTCTTTCCCTACACGACGCTCTTCCGATCTGGGCCACCTCCAGTGC | Illumina I5 Primer |
| ag1651 | AATGATACGGCGACCACCGAGATCTACACCACCGATGTGACACTCTTTCCCTACACGACGCTCTTCCGATCTGGGCCACCTCCAGTGC | Illumina I5 Primer |
| ag1652 | AATGATACGGCGACCACCGAGATCTACACTGTTAGCACAACACTCTTTCCCTACACGACGCTCTTCCGATCTGGGCCACCTCCAGTGC | Illumina I5 Primer |
| ag1653 | AATGATACGGCGACCACCGAGATCTACACTTGGCAGCGTACACTCTTTCCCTACACGACGCTCTTCCGATCTGGGCCACCTCCAGTGC | Illumina I5 Primer |
| ag1654 | AATGATACGGCGACCACCGAGATCTACACCCAATGATACACACTCTTTCCCTACACGACGCTCTTCCGATCTGGGCCACCTCCAGTGC | Illumina I5 Primer |
| ag1371 | ATTAGGCGCGCCGCGGCCGCNNNNNNNNNNNNNNNNNNGTCGACCAATTGATTA | pAG424 Barcode F |
| ag1372 | TAATCAATTGGTCGAC | pAG424 Barcode R |
| ag491 | TTCAGCAATTGGCACTGGAGGTGGCCC | Jain F pET01 barcode RE site |
| ag492 | GGTGGCGCGCCTCCACCCAGCTCCAGTTGT | Jain R pET01 barcode RE site |
| mo38 | GATCCACGATGC | pAG424 gene specific RT primer |
| mo37 | GGATTCTTCTACACACCC | pAG424 sequencing primer F |
| ag489 | GGGCCACCTCCAGTGC | pAG424 sequencing primer R |
| ag490 | CTGGATTGTCCTATGTGTCTTTGCTTCT | pAG424 barcode sequencing primer F |
| mo367 | CCCAGGGTGCGGTGAGATCCCAGAGGA | SCN5A QC SP T1131I |
| ag1749 | TTGACAACGTGGGGGGCGGGTACCTGGC | SCN5A QC SP A1407G |
| mo346 | CACCGTTGCACTCAGACCACGCCAT | c.1891-5C>G Guide F |
| mo347 | AAACATGGCGTGGTCTGAGTGCAAC | c.1891-5C>G Guide R |
| mo348 | TCGAAGCCATCTACACACGGAGCCTGGGAGGTCAGCATCTGGGGCCCGCCTGGCTCCTCTGATGGCGTGGTCTGACTGCAATCAGGAGATTTGCGTCAGCCTGGGGAAAAGGGTCCTGCCCCCAGCTCCTGTCCTGCTGGACCTGGGGAGG | c.1891-5C>G repair template with PAM break |
| mo198 | GGAGGACCTGTGATATGTGTAGC | c.1891-5G>A genotyping F |
| mo199 | AGATAGACATGGTTATGGTTGGGA | c.1891-5G>A genotyping R |

QC – QuikChange; SP – SyncroPatch.

**Supplemental Table II.** Construct exon and flanking intron lengths.

| **Construct** | **Exon BPs** | **Upstream BPs** | **Downstream BPs** |
| --- | --- | --- | --- |
| 5 | 129 | 250 | 250 |
| 6 | 92 | 100 | 152 |
| 7 | 231 | 250 | 250 |
| 8 | 64 | 250 | 250 |
| 9 | 142 | 250 | 250 |
| 10 | 198 | 125 | 125 |
| 11 | 180 | 250 | 250 |
| 12 | 372 | 125 | 125 |
| 13 | 133 | 250 | 250 |
| 14 | 239 | 250 | 250 |
| 16 | 351 | 250 | 250 |
| 18 | 162 | 250 | 250 |
| 19 | 121 | 250 | 250 |
| 20 | 155 | 250 | 250 |
| 21 | 174 | 250 | 250 |
| 22 | 123 | 250 | 250 |
| 23 | 282 | 250 | 250 |
| 24 | 54 | 250 | 250 |
| 27 | 271 | 250 | 250 |
